# Proposed Mammography Protocol and Pre-Mammography Survey for Transgender Patients

**DOI:** 10.1101/2024.06.13.24308890

**Authors:** Carlos Carrasco Godoy, Diego Muñoz Fernández, Sergio Quitral Urzúa, Elizabeth Cañipa Zulueta, Marco Jiménez Herrera, Jeannette Soto Miranda

## Abstract

**Background:** Early detection of breast cancer reduces the risk of mortality, with mammography being the gold standard for detecting breast cancer. Currently, there is no consensus on breast screening in transgender patients, with only recommendations from various international health organizations, which differ from one another. The objective of this research was to determine the mammographic projections applicable to transgender patients, in order to propose a mammography protocol and a pre-mammography survey focused on this patient group.

**Methodology:** A semi-structured survey with both closed and open-ended questions was conducted, which was answered by radiologists specializing in mammography. To propose mammographic projections, the survey included questions about the transitional phase, hormone use, surgeries, and breast tissue development. To propose a pre-mammography survey for transgender patients, questions about risk factors and surgical treatments were included.

**Conclusions:** A mammographic protocol is proposed according to the transitional phase, along with a pre-mammography survey for transgender patients, based on information gathered from a survey answered by 20 radiologists specializing in mammography. This contributes significantly to the screening process for this patient group.

## Background

Breast cancer is the most common neoplasm in women worldwide, and in 2018, approximately 2 million new cases were diagnosed, making it the leading cause of cancer death among women globally(Nardin et al., 2020). This disease is characterized by the formation of malignant tumors originating from breast cells, which can invade surrounding tissues and metastasize to other parts of the body (Harbeck et al., 2019).

Despite significant advances in the detection and treatment of breast cancer, which have improved survival rates, the incidence of this disease continues to increase globally. It is essential to continue researching new therapeutic strategies and prognostic factors to further improve patient management and outcomes (Smolarz et al., 2022).

Although it is much more common in women, men can also develop breast cancer, albeit less frequently (Veronesi et al., 2005). However, there is another population that also faces a risk of developing breast cancer: transgender individuals. Transsexuality is a term commonly used to describe both individuals who undergo medical procedures to modify their physical appearance according to their gender identity and those who do not make such a decision. Therefore, the term “trans person” encompasses transgender individuals, transsexuals, and cross-dressers.

Transfeminine individuals are biologically male-bodied individuals who dress, feel, and identify as female, while transmasculine individuals are biologically female-bodied individuals who dress, feel, and identify as male (Safer & Tangpricha, 2019).

Transgender men who have undergone surgery have a lower risk of breast cancer compared to cisgender women (Stone et al., 2018), On the other hand, transgender women receiving feminizing hormone therapy (FHT) have a higher risk of breast cancer compared to cisgender men, but lower than that of cisgender women (Banks et al., 2003; De Blok et al., 2019; Lagro-Janssen et al., 2003).

To evaluate the breast for both screening and diagnosis, mammography is used with two routine projections: Cranio-caudal (CC) and Medio-lateral oblique (MLO) (Vujovich et al., 2018). These projections are standardized for the cisgender population, but there is limited literature studying or proposing appropriate projections for transgender patients.

It is suggested that transgender women follow screening guidelines for cisgender women if they have risk factors and have been on hormone therapy for more than 5 years. For transgender men, the risk of breast cancer significantly decreases after bilateral mastectomy, but it is recommended to follow screening guidelines for cisgender women if breast tissue is retained (Chowdhry & O’Connell, 2020a; Parikh et al., 2020).

Given these considerations, there is currently no clear consensus or defined protocol regarding whether conventional projections are adequate for transgender patients. Previous studies have only provided recommendations from various authors as mentioned earlier. Therefore, the purpose of this study is to determine the appropriate mammographic projections for transgender patients. Ultimately, this aims to propose a mammography protocol specifically tailored to this patient group and make recommendations on the necessary mammographic projections based on the health characteristics of transgender individuals, whether they are transgender men or women.

## Materials and Methods

This study has been approved by the ethics committee of our institution.

This observational, descriptive, cross-sectional study collected data from 20 radiologists through a survey validated by two experts in the field with over 20 years of experience (validation in supplementary material). The instrument used was a semi-structured questionnaire containing both closed and open-ended questions for data collection, with responses measured at a nominal level. The survey was administered via Google Forms.

At the outset of the survey, participants were asked for their consent to participate. Following this, a brief context and definitions regarding the transgender population, breast cancer associated with this population, and the transition experience were provided. This aimed to furnish the minimal necessary information to answer subsequent questions, enabling non-expert professionals to participate actively in the survey.

Closed-ended questions contained predefined response categories or alternatives. Participants were presented with possible responses and required to select the option that best described their answer. Closed questions could be dichotomous (two response alternatives) or include multiple response options defined a priori by the researcher. Some closed questions allowed participants to select more than one response option. Conversely, open-ended questions did not predefine response alternatives.

Numeric values were assigned to data as needed for quantitative analysis. Open-ended responses were coded once all participant responses were collected, or at least after identifying major response trends in a sample of the administered questionnaires. The procedure involved identifying and naming general response patterns (similar or common responses), listing these patterns, and then assigning a numeric value or symbol to each pattern, thereby constituting response categories. The following procedure was used to finalize open-ended questions:

1. Selecting the number of responses using an appropriate sampling method to ensure the representativeness of the surveyed participants.
2. Observing the frequency with which each response appears in the questions.
3. Choosing the responses that appear most frequently.
4. Classifying the responses according to a logical criterion, ensuring they are mutually exclusive.
5. Titling each theme in a representative manner.
6. Assigning a code to each general response pattern.

A total of 10 closed-ended questions were conducted, with an open-ended question added respectively from question number 1 to number 10, allowing radiologists to provide additional information on mammographic projections not mentioned or any comments they deem useful for the mammography technique protocol for transgender patient care. A coherence table (supplementary material) was developed to facilitate the interpretation of results afterward. Additionally, the final survey proposal prior to mammography was adapted from a survey for cisgender patients conducted by (Muñoz Villaseca et al., 2024). Part of these researchers are involved in both studies.

For data analysis, responses were coded numerically and subsequently transformed into percentages to determine which responses were most frequent. This approach was used to develop the protocol proposal for transgender patients.

## Results

Below is each question with its respective results, which were answered by 20 radiologists specializing in mammography who agreed to respond to each survey question.

To determine specific mammographic projections for transgender women based on their transition phase, the following questions were asked:

1. Question No. 1 - What mammographic projection would you request for a transgender woman over 40 years old who has been on hormonal treatment for 5 years or more? 100% find the medio-lateral oblique projection useful. 80% find the cranio-caudal projection useful. 30% find breast ultrasound useful. Regarding open-ended question No. 1, would you consider performing another mammographic projection? 70% do not consider it necessary to perform another mammographic projection. 30% suggest performing additional projections, such as magnification and axillary projection, but only in case of findings in the routine CC and MLO projections.
2. Question No. 2 - What mammographic projection would you request for a transgender woman over 40 years old, whose hormonal treatment has been for less than 5 years? 70% find the medio-lateral oblique projection useful. 55% find the cranio-caudal projection useful. 35% find breast ultrasound useful. 2% consider mammography unnecessary. Regarding open-ended question No. 2, would you consider performing another mammographic projection? 55% do not consider it necessary to perform an additional mammographic projection. 25% suggest performing additional projections, such as magnification and axillary projection, but only in case of findings in the routine CC and MLO projections. 20% did not respond to the question.
3. Question No. 3 - What mammographic projection would you request for a transgender woman over 40 years old who is not on hormonal treatment? 70% consider mammography unnecessary. 15% consider requesting a breast ultrasound necessary. 15% consider requesting a medio-lateral oblique projection necessary. 2% consider requesting a cranio-caudal projection necessary.
4. Question No. 4 - What mammographic projection would you request for a transgender woman over 40 years old who has breast tissue due to hormonal treatment and also has breast implants? 100% consider cranio-caudal and medio-lateral oblique projections necessary. 85% consider the Eklund maneuver necessary. 35% consider requesting a breast ultrasound necessary.
5. Question No. 5 - What mammographic projection would you request for a transgender woman over 40 years old who has not developed breast tissue and only has breast implants? 30% consider requesting a cranio-caudal projection necessary. 35% consider requesting a medio-lateral oblique projection necessary. 25% consider the Eklund maneuver necessary. 70% consider requesting a breast ultrasound necessary. 15% do not consider mammography necessary. Regarding transgender female patients, regarding additional responses from open-ended questions No. 3, No. 4, and No. 5, regarding whether to add another mammographic projection, 100% of respondents answered that it is not necessary to perform another mammographic projection, respectively. To determine specific mammographic projections for transgender men based on their transition phase, the following questions were asked:
6. Question No. 6 - What mammographic projection would you request for a transgender man over 40 years old who has not yet undergone bilateral mastectomy (i.e., still retains breasts)? 100% consider cranio-caudal and medio-lateral oblique projections necessary. 20% consider requesting a breast ultrasound necessary.
7. Question No. 7 - What mammographic projection would you request for a transgender man over 40 years old who has already undergone bilateral mastectomy? 10% consider requesting a cranio-caudal projection necessary. 10% consider requesting a medio-lateral oblique projection necessary. 50% do not consider mammography necessary. 60% consider requesting a breast ultrasound necessary.
8. Question No. 8 - What mammographic projection would you request for a transgender man over 40 years old who has undergone only breast reduction surgery? 90% consider cranio-caudal and medio-lateral oblique projections necessary. 35% consider requesting a breast ultrasound necessary. Regarding transgender male patients, regarding additional responses from open-ended questions No. 6, No. 7, and No. 8, regarding whether to add another mammographic projection, 100% of respondents answered that it is not necessary to perform another mammographic projection, respectively. To identify the obstacles faced by radiologists specializing in mammography during the diagnostic evaluation process of transgender patients, the following questions were asked:
9. Question No. 9 - What obstacles do you encounter when generating a radiological report for a transgender patient? 10% indicate that the ACR breast classification time is an obstacle for transgender patients. 10% indicate that transgender patient risk factors are a hindrance. 35% indicate insufficient knowledge about LGBTIQ+ terminology. 60% indicate insufficient experience in caring for transgender patients. 25% indicate that the presence of gynecomastia in transgender patients is an obstacle. Regarding open-ended question No. 9, do you consider any other significant obstacles? 80% state that they do not have any other obstacles to add. 10% mention having insufficient knowledge on the topic. 5% mention that obtaining medical history from transgender patients is a hindrance. 5% mention that transgender patient hormone use duration is a hindrance. To propose a mammography protocol and questionnaire for the care of transgender patients, the following questions were asked:
10. Which of the following medical histories do you believe is necessary to include in the mammography questionnaire for proper management of a transgender patient? 85% believe it is necessary to include the patient’s biological sex. 75% believe it is necessary to include the patient’s gender identity. 60% believe it is necessary to include whether the patient has undergone gender reassignment surgeries. 100% believe it is necessary to include whether the patient has undergone breast surgeries. 100% believe it is necessary to include whether the patient is undergoing hormone replacement therapy and the duration of use. Regarding open-ended question No. 10, do you consider any other important medical history? If so, which one(s)? 50% believe it is not necessary to add another important medical history to the mammography questionnaire. 25% believe it is important to include the patient’s family history of breast cancer in the mammography questionnaire. 10% believe it is necessary to include the type of hormone treatment in the mammography questionnaire. 5% believe it is important to include menarche in the mammography questionnaire, as well as whether the patient has children or has breastfed, specifically for transgender men. 10% did not respond to the question.

Based on the responses provided by surveyed radiologist specialists in mammography, specific mammographic projections for transgender patients were determined depending on their phase of transition. This is presented as a proposed mammography protocol for the care of transgender patients in Tables 1 and 2.

**Table 1.** Transgender Women.

| Pase | Mammographic Projection Applicable |
| --- | --- |
| >40 years and HRT ≥ 5 años | CC y MLO Bilateral |
| >40 years and HRT < 5 años | CC y MLO Bilateral |
| >40 years and without HRT | No mammogram needed |
| <40 years, with breast tissue development and implants | CC, MLO and bilateral Eklund maneuver |
| <40 years, with breast tissue development and implants | Bilateral breast ultrasound |

HRT: hormonal replacement therapy

**Table 2.** Transgender Men.

| Phase | Mammographic Projection Applicable |
| --- | --- |
| >40 years and without mastectomy | CC y MLO Bilateral |
| >40 years with mastectomy | Bilateral breast ultrasound |
| <40 years without breast reduction | CC y MLO Bilateral |
| <40 years with breast reduction | Bilateral breast ultrasound |

Note:
Initiate screening before the age of 40 if the transgender patient has a family history of breast cancer or carries BRCA1/BRCA2 genetic mutations, as these factors increase the likelihood of developing breast cancer.

Tables 1 and 2 - Mammographic projections applicable to transgender patients according to their phase of transition as a proposed mammography protocol.

The final mammography survey for transgender patients was supplemented and adapted from the survey conducted by (Muñoz Villaseca et al., 2024) resulting in the following model to be delivered to patients prior to undergoing mammography.

### Mammography Survey Proposal

Full Name:

ID Number:

Age:

Phone:

Reason for Consultation: Date of Exam:

BIOLOGICAL SEX

**□ Male**

**□ Female**

### GENDER WITH WHICH YOU IDENTIFY

**□ Male**

**□ Female**

**Previous exams**

| Exam | Date |
| --- | --- |
| Mx |  |
| Ultrasound |  |
| MRI |  |

**Signs and symptoms**

|  | Breast<br>Right | Breast<br>Left | Date |
| --- | --- | --- | --- |
| Asymptomatic |  |  |  |
| Pain |  |  |  |
| Lump |  |  |  |
| Skin Engorgement |  |  |  |
| Skin Retraction |  |  |  |
| Discharge |  |  |  |
| Inverted Nipple |  |  |  |
| Other: |  |  |  |

Observations:

### Family history of breast cancer

| Relative | Paternal | Maternal | Years |
| --- | --- | --- | --- |
| Grandmother |  |  |  |
| Mother |  |  |  |
| Aunt |  |  |  |
| Sister |  |  |  |
| Daughter |  |  |  |
| Other: |  |  |  |

### Surgical history

|  | Right breast | Left breast | Date |
| --- | --- | --- | --- |
| Mastectomy (Indicate if it is total or partial) |  |  |  |
| Biopsy (Indicate if it is stereotaxic, excisional or core) |  |  |  |
| Augmentation |  |  |  |
| Reduction |  |  |  |
| Radiotherapy |  |  |  |
| Chemotherapy |  |  |  |
| Mastitis |  |  |  |
| Hormone Replacement Therapy |  |  |  |
| Treatment Duration |  |  |  |
| Other |  |  |  |

### Gynecological-obstetrical history

Age of menarche:

Menopause (indicate age and whether it was natural or surgical):

Number of children:

Age first pregnancy:

Lactation:

Oophorectomy:

Currently pregnant:

Tamoxifen consumption:

### Modifiable factors

Alcoholism:

Smoking:

Weight and size:

## Discussion and Conclusions

After collecting data from 20 specialist radiologists in breast imaging, the mammography survey is proposed, including valuable information on transgender patient histories. Additionally, mammographic projections are recommended based on these patients’ characteristics. The differentiation between transgender men and women becomes crucial, as described by (Stone et al., 2018), since risks decrease or increase depending on the patient’s condition. Therefore, this survey makes the distinction and classifies according to the patient’s condition.

The mammography survey sent to radiologists is an unprecedented compilation of information, as most of the reviewed evidence recommends the same projections for cisgender patients (Chowdhry & O’Connell, 2020a), and are not included in early detection guidelines. However, we believe it is valuable, as we confirmed with these 20 respondents, that the projections and imaging modality will depend on whether the transgender person is male or female, their age, whether they have had mastectomy, breast implants, and the hormonal treatment they may be undergoing (see Tables 1 and 2). Therefore, the recommended radiographic projections are as follows: For a transgender woman over 40 years old who has undergone hormonal treatment for 5 years or more, it is recommended to perform mammography with Craniocaudal and Mediolateral Oblique projections, as also recommended (Chowdhry & O’Connell, 2020b; Parikh et al., 2020). by the guidelines. If an obvious finding is detected, additional projections should be performed; otherwise, if no findings are present, further irradiation is unnecessary. For a transgender woman over 40 years old whose hormonal treatment has been for less than 5 years, it is also recommended to perform mammography with routine Craniocaudal and Mediolateral Oblique projections, as concluded (De Blok et al., 2019), if no significant physiological changes are evident, a breast ultrasound should be performed. If an obvious finding is detected, additional projections should be performed; otherwise, if no findings are present, further projections are unnecessary. For a transgender woman over 40 years old not undergoing hormonal treatment, it is recommended not to perform mammography (in the absence of family history and signs of breast abnormalities). For a transgender woman over 40 years old with breast tissue development due to hormonal treatment and breast implants, it is recommended to perform mammography with Craniocaudal and Mediolateral Oblique projections, complemented by the Eklund maneuver. For a transgender woman over 40 years old who has not developed breast tissue but has breast implants, mammography is not necessary; bilateral breast ultrasound is sufficient. For a transgender man over 40 years old who has not undergone bilateral mastectomy, mammography with routine Craniocaudal and Mediolateral Oblique projections is recommended. For a transgender man over 40 years old who has already undergone bilateral mastectomy, mammography is unnecessary; only bilateral breast ultrasound is recommended. For a transgender man over 40 years old who has undergone breast reduction surgery, mammography with routine Craniocaudal and Mediolateral Oblique projections is recommended.

One of the drawbacks of this study is that although our initial target sample size was forty specialist radiologists in mammography from Chile, we only received responses from twenty radiologists, and the reasons for non-response from the remaining radiologists are unknown. However, we consider the data from 20 radiologists to be sufficient and highly valuable. Another limitation concerns whether all surveyed radiologists have experience in providing care and reporting mammograms for transgender patients.

Currently, there is limited scientific information on breast cancer in the transgender population due to the limited number of studies addressing this topic. The main advantage of this study is that it proposes recommendations for a population that has been understudied, based on data from 20 expert radiologists in mammography. Given the sparse existing literature, this study substantially contributes to knowledge and the development of protocols aimed at more informed diagnosis.

The findings of this study aim to gather more evidence to complement the existing knowledge and encourage further studies focused on early diagnosis, educating both the transgender community and radiologists interpreting mammographic images.

## Data Availability

All data produced in the present study are available upon reasonable request to the authors

